# Li-Fraumeni Syndrome-Associated p53 Variants Disrupt Kidney and Urinary Tract Development

**DOI:** 10.1101/2025.08.29.25333303

**Authors:** Adrian Romero, Alejandra Davilavaladez, Alexandria T. M. Blackburn, Amisheila Kinua, Yogesh Srivastava, D. Gareth Evans, Elizabeth K. Bancroft, Rosalind A. Eeles, Mark E. Corkins, Rachel K. Miller

## Abstract

Li-Fraumeni Syndrome (LFS) is a rare autosomal dominant disorder that increases the risk of various types of cancer. It is primarily caused by inherited mutations in the *TP53* gene. While the tumor suppressor function of p53 is well established, its role in embryonic development, particularly in the formation of the kidney and urinary tract, remains poorly understood. Moreover, its contribution to human congenital anomalies has not been clearly defined. Here, we report that pathogenic *TP53* variants can lead to congenital anomalies of the kidney and urinary tract (CAKUT), as well as genital defects (GD), in individuals with LFS. Among 28 unrelated *TP53* mutation carriers, 28% (8/28) exhibited CAKUT and/or GD, with a higher frequency observed in individuals carrying structurally disruptive or dominant-negative mutations. We focused on two clinically observed variants: R242W, which destabilizes protein structure, and R282W, a dominant-negative hotspot mutation. AlphaFold modeling showed that both variants cluster within the DNA-binding domain and are predicted to disrupt tetramer formation. In *Xenopus laevis, tp53* is expressed in developing nephric structures, consistent with findings from mouse models of nephrogenesis. Expression of either mutant *TP53* mRNA in *Xenopus embryos* disrupted kidney morphogenesis *in vivo*, supporting a developmental loss-of-function effect. These findings indicate that pathogenic *TP53* variants contribute to renal and urogenital defects in LFS. They reveal a previously unrecognized developmental role for p53 and expand the phenotypic spectrum associated with this cancer predisposition syndrome.

## Introduction

The tumor suppressor gene *TP53*, encoding the p53 protein, is widely recognized as the “guardian of the genome” for its critical role in maintaining genomic stability (Lane, 1992, Marei et al., 2021). As a transcription factor, p53 induces genes involved in DNA repair, cell cycle arrest, and apoptosis in response to cellular stress (Kuerbitz et al., 1992, Smith et al., 1994, Miyashita and Reed, 1995, el-Deiry et al., 1993). Because p53 functions as a tetramer, many *TP53* mutations can exhibit dominant-negative effects, often leading to early-onset cancer predisposition syndromes called Li-Fraumeni syndrome (LFS) (Monti et al., 2011, Friedman et al., 1993, Gaglia et al., 2013). Beyond its well-known tumor-suppressive role, p53 also plays essential, yet often overlooked, functions in embryonic development and organogenesis (Saifudeen et al., 2009).

Although p53-null mice are viable, they develop tumors early in life (Donehower et al., 1992), and certain genetic backgrounds reveal developmental defects, including neural tube closure abnormalities and craniofacial malformations (Armstrong et al., 1995, Rinon et al., 2011). Additionally, a subset of p53-deficient mice displays growth retardation or subtle developmental anomalies involving the palate, eyes, teeth, lungs, or kidneys (Kaufman et al., 1997, Saifudeen et al., 2009, Baatout et al., 2002). Notably, renal hypoplasia and duplex ureters have been observed in p53-deficient mice, implicating p53 in nephrogenesis (Saifudeen et al., 2002, Saifudeen et al., 2009). Conditional deletion of *Trp53* in nephron progenitor cells results in the loss of cap mesenchyme, a key population required for nephron formation (Li et al., 2015). Complementary studies in *Xenopus laevis* show that dominant-negative p53 overexpression leads to pronephric loss, further supporting p53’s conserved role in kidney development (Wallingford et al., 1997). Mice expressing a stabilized, transcriptionally inactive *Trp53* variant show embryonic lethality and multiple congenital malformations, including craniofacial, ocular, and cardiac anomalies (Van Nostrand et al., 2014). On the other hand, overexpression of overactive p53 also causes tissue-specific developmental defects. These phenotypes are likely due to inappropriate activation of p53-mediated apoptosis or proliferation arrest during critical stages of organogenesis (Bowen and Attardi, 2019).

While p53 has been implicated in kidney development in animal models, its contribution to congenital anomalies of the kidney and urinary tract (CAKUT) in humans has not been directly demonstrated. In this study, we investigated the potential developmental role of *TP53* variants identified in LFS patients presenting with CAKUT and/or GD abnormalities. We used *Xenopus laevis*, a well-established model for studying pronephric kidney development, due to its accessible embryos, rapid kidney organogenesis, and structural and molecular similarity to the mammalian nephron (Blackburn and Miller, 2019, Zhou and Vize, 2004, Corkins et al., 2023). Using unilateral microinjection approaches, we examined the effects of two patient-derived *TP53* variants on kidney development. Our findings demonstrate that both variants disrupt pronephric morphogenesis in *Xenopus* and are associated with CAKUT in human patients, providing evidence that p53 is essential for kidney development.

## Materials and methods

### Study participants and data analysis

Human *TP53* mutation carriers as part of the SIGNIFY study (Saya et al., 2017) were scanned with Diffusion-Weighted (DW) whole body MRI from November 2012 through July 201. Individuals carrying known low-penetrance *TP53* mutations (as determined by a clinical geneticist) or variants of uncertain significance were excluded, as were those with a malignancy diagnosed within the previous five years, except for non-melanoma skin cancer or cervical carcinoma *in situ* (CIS). For this paper, an additional sub-study involving 44 patients was conducted to examine renal morphology and function, with data collected on renal function, body mass index (BMI), and blood pressure. However, only 28 patients were included in the substudy assessing blood pressure and glomerular filtration rate (GFR). This component was added later to the protocol. Participants who lived locally were recalled for these follow-up measurements; for others, participation was not feasible due to logistical constraints. Consequently, inclusion in this substudy was determined solely by the feasibility of returning for additional evaluation, without any selection or prioritization based on clinical characteristics. The research was approved by the Health Research Authority NRES Committee London—Brent (12/LO/0781).

### *TP53* Testing and Sequencing

The entire *TP53* coding region (exons 2–11) was analyzed by direct sequencing, with a reported sensitivity of >98%. Negative results do not exclude somatic or germline mosaicism. Deletions and duplications were assessed using Multiplex Ligation-dependent Probe Amplification (MLPA, MRC-Holland kit P056). Single exon deletions/duplications were confirmed by sequencing and/or dosage-sensitive WAWE analysis. Mutation nomenclature follows Human Genome Variation Society (HGVS) guidelines, with numbering based on the A of the ATG start codon (reference sequence: GenBank NM_000546). Sequencing was performed using the Illumina TruSight Cancer Panel, analyzed via GAMA and SIGMA bioinformatic pipelines. The full coding sequence and intron-exon boundaries were screened for small variants. Pathogenic and rare variants were confirmed by bidirectional Sanger sequencing.

### MRIs

MRI examinations were performed at The Royal Marsden NHS Foundation Trust or Central Manchester University Hospitals NHS Foundation Trust using a 1.5 T MRI machine (Siemens, Erlangen, Germany) from vertex to feet using conventional MR imaging sequences (T1-weighted, with or without fat suppressed T2-weighted/STIR sequences), as well as diffusion weighted images. Slices were 0.8 cm thick and scans were not contrast enhanced.

Scans were read independently by two radiologists with at least 5 years’ experience, who were blinded to the mutation status of the study participants. Ten percent of scans, selected randomly, were cross-read at both centres for quality assurance. Data sheets were completed for each participant to collate clinical data, including blood pressure, BMI and renal function.

### *Xenopus laevis* embryos and microinjections

*Xenopus* eggs were obtained using standard procedures, incubated in 0.3x MMR, and fertilized *in vitro* (Sive et al., 2012). Blastula cleavage stages and dorsal-ventral polarity were identified using established methods (Nieuwkoop and Faber, 1994). Microinjections were specifically targeted to the V2 blastomere at the eight-cell stage, as this lineage significantly contributes to pronephros development (Nieuwkoop and Faber, 1994, DeLay et al., 2016, Moody and Kline, 1990). For the overexpression of wild-type human *TP53* (p53 WT) and its mutants (*TP53*^*R248W*^ or *TP53*^*R282W*^), a mixture containing 50 pg of *TP53* mRNA and 300 pg of membrane-RFP mRNA (Davidson et al., 2006) was injected as a lineage tracer to confirm the targeted blastomere. As a control, 50 pg of β-galactosidase mRNA was co-injected with the tracer. Details on experimental design and statistical analysis are provided in Supplementary Information (Table S1). All procedures were approved by the UTHealth Center for Laboratory Animal Medicine Animal Welfare Committee (IACUC protocol #: AWC-22-0082).

### Whole-mount *in situ* hybridization

A digoxigenin-11-UTP-labeled antisense RNA probe for *tp53* was synthesized by in vitro transcription using T7 RNA polymerase. The DNA template was generated through PCR amplification from plasmid 415-AP, obtained from the European *Xenopus* Resource Centre (EXRC) (XB-CLONE-12440929) (Wallingford et al., 1997). This clone contains the *Xenopus laevis* coding sequence for *tp53*. The primers used to generate the template are forward CTAGCTAATACGACTCACTATAGGGCTCTGGGGTCTCGAGGGTGA and reverse CCCACCGTCACTTCATGTGC. The embryos were staged, fixed, and stained according to standard procedures (Nieuwkoop and Faber, 1994, Sive et al., 2012) using an anti-digoxigenin antibody (1:3000, Sigma 11093274910, St. Louis, MO, USA) and BM Purple (Sigma 11442074001).

### Hybridization Chain Reaction (HCR)

For *in situ* Hybridization Chain Reaction (HCR) on whole *Xenopus laevis* embryos, staging and fixation were performed according to standard protocols (Nieuwkoop and Faber, 1994, Sive et al., 2012), followed by a modified *in situ* HCR protocol based on (Choi et al., 2018, Schock et al., 2024). Hybridization probes for *lhx1* and *atp1a1* were designed and synthesized by Molecular Instruments Inc. (https://www.molecularinstruments.com/), which also provided the probe hybridization buffer, probe wash buffer, amplification buffers, and DNA HCR amplifier hairpin sets. The HCR probe for *tp53* was designed using a probe maker tool developed by Ryan Null (Kuehn et al., 2022) and executed via Anaconda Navigator (2023-07-1) and JupyterLab (3.0.10). Five embryos were collected in Wheaton vials (4 mL) and fixed in 4% formaldehyde in 1× MEMFA (MOPS, EGTA, MgSO_4_) for 60 min at room temperature (RT) on a rotator. After fixation, embryos were washed three times (5 min each) with PBST, followed by three washes (5 min each) with 100% methanol, and permeabilized overnight at − 20°C in 100% methanol. The next day, embryos were rehydrated through sequential 5-min washes in 75% methanol/25% PBST, 50% methanol/50% PBST, and 25% methanol/75% PBST, followed by two washes in 100% PBST (5 min each). Embryos were then washed twice in 0.1 M triethanolamine (5 min each). Acetylation was performed by sequential 5-min incubations in 0.1 M triethanolamine containing 0.25% acetic anhydride, followed by 0.5% acetic anhydride. After two additional PBST washes (5 min each), embryos were refixed in MEMFA for 20 min at RT on a rotator and then washed five times in PBST (5 min each). Embryos were incubated in preheated (37°C) wash buffer (500 μL) for 5 min, then transferred to hybridization buffer (500 μL) at 37°C for 30 min. The buffer was replaced with probe solution (8 nM probe diluted in hybridization buffer), and embryos were incubated overnight (12–16 hrs) at 37°C. After hybridization, embryos were washed four times (15 min each) in probe wash buffer at 37°C. For amplification, embryos were incubated in 500 μL of amplification buffer for 30 min at RT, then transferred to an amplification solution containing fluorescently tagged hairpin pairs (h1 and h2). Hairpins were preheated to 95°C, incubated for 30 min at RT in the dark, diluted in amplification buffer (final concentration: 48 nM), and added to the embryos for overnight incubation (12–16 hrs) at RT in the dark. The following day, embryos were sequentially washed in 5× SSCT (two 5-min washes, one 30-min wash), followed by a 30-min wash in 5× SSCT containing DAPI. After three PBST washes (5 min each) in the dark and three PBS washes (5 min each) in the dark. Embryos were preserved in PBS for immediate imaging or in methanol for long-term storage and imaging.

### Western blot

Ten *Xenopus* embryos were collected at various stages to prepare protein lysates as previously described (Kim et al., 2002). One embryo equivalent of lysate was loaded per well on a 10% SDS-PAGE gel. Proteins were transferred to 0.45 μm nitrocellulose membranes and blocked for 1 hour at room temperature in KPL block (SeraCare) at 4°C. Membranes were incubated overnight at 4°C with primary antibodies: mouse anti-p53 (1:1000, Abcam ab16465), mouse anti-Flag (1:1000, Sigma M2), or rabbit anti-GAPDH (1:1000, Santa Cruz). After washing with TBS-T, membranes were incubated for 1 hour at room temperature with goat anti-rabbit or goat anti-mouse IgG horseradish peroxidase-conjugated secondary antibody (1:3000; Bio-Rad). Following further washing with TBS-T, blots were imaged using the Bio-Rad ChemiDoc XRS+ system and SuperSignal West Pico PLUS Chemiluminescent Substrate (Thermo Fisher Scientific).

### Imaging

Embryos used for *in situ* hybridization were imaged on a Zeiss AxioZoom V16 with a 1x objective, Zeiss 512 color camera (Zeiss, Oberkochen, Germany), and extended depth of focus processing. Embryos used for HCR were scored and photographed using a Zeiss LSM800 confocal microscope (Zeiss, Oberkochen, Germany). HCR-stained embryos were dehydrated in 100% methanol and cleared in BABB/Murray’s 1:2 (volume of benzyl alcohol to benzyl benzoate) solution for 10 min. Embryos were mounted on cover glasses and imaged using an LSM800 inverted confocal microscope. Image processing, including maximum intensity projection of z-stacks and contrast adjustments, was performed using Imaris software.

### AlphaFold analysis

The DNA sequence used for modeling the p53 tetramer binding site was adopted from (McLure and Lee, 1998). AlphaFold 3 was used for structural modeling of the p53-DNA complex (for clear visualization of binding, DNA extended from both ends by adding 7 A-T nucleotides), which enabled precise prediction of the interactions between the DNA-binding domain and the p53 tetrameric interface. The AMBER ff14SB force field for proteins and AMBER Bsc0 force field for nucleic acids were used to energy minimize for the final models. R248Q and R282W mutations were introduced by in silico site-directed mutagenesis using the swapaa command in order to examine the structural impact of mutations linked to cancer. With a step size of 0.02 Å, a two-stage procedure including 1,000 steps of the steepest descent method and 500 steps of the conjugate gradient algorithm was used to minimize energy. UCSF ChimeraX was used for distance measurements and molecular visualization. The stability of the tetrameric interface, residue interactions with DNA, and possible steric conflicts brought on by mutations were all evaluated structurally. The sequences used for AlphaFold analysis were downloaded from the NIH GenBank repository: NM_000546_6 Homo sapiens tumor protein p53 (TP53), transcript variant 1, mRNA.

## Results

### CAKUT/GD has been identified in patients with *TP53* variants, most of which are loss-of-function mutations located in the DNA-binding domain (DBD) of p53

A total of 88 participants were recruited from 37 families (Table 1), including 44 carriers of pathogenic *TP53* variants and 44 matched healthy population controls. Among the carriers, 18 participants had a prior cancer diagnosis, and 6 of these individuals had experienced multiple primary tumors. During the study period, 6 carriers were diagnosed with a malignancy, 4 of which were detected directly through whole-body MRI screening. No cancers were identified in the control group.

**Table 1:** Characteristics of the cases and controls with previous malignant tumours.

|  | Carriers | Controls |
| --- | --- | --- |
| N | 44 | 44 |
| Age, median (range) | 38 (19-58) | 38 (22-59) |
| Female, n (%) | 27 (61%) | 27 (61%) |
| Male, n (%) | 17 (39%) | 17 (39%) |

MRI data revealed that a cohort of Li-Fraumeni patients from 19 to 58 years of age exhibited a higher prevalence of CAKUT relative to the control group. CAKUT manifested itself in kidney hypoplasia, complete agenesis of the kidneys and reproductive tract, renal cysts, benign kidney tumors, ovarian cysts, and reduced glomerular filtration rates (Table 1). 57 % (16/28) of the patients exhibited either structural urogenital anomalies and/or abnormal glomerular filtration rates (GFRs) below 90 mL/min/1.73 m^4^ (normal range: 90–120 mL/min/1.73 m^4^) (Table 2). Of the 57% of patients with p53 variants who exhibit genitourinary (GU) abnormalities, eight present with structural defects observable by MRI. Several patients have notable anomalies such as simple renal cysts, renal angiomyolipoma, a large adenoma, and complex cysts. These findings highlight a spectrum of kidney defects, ranging from mild to clinically significant, supporting a critical role for p53 in GU development. Additionally, eight patients without obvious structural anomalies exhibit a glomerular filtration rate (GFR) below 90, suggesting impaired kidney function despite a normal anatomy (Table 2).

**Table 2.** Demographics and molecular data of patients with *TP53* SNVs and small indels (<10bp)

| Patient ID | Sex | Exon | Nucleotide change | AA change | PD | Effect | SIFT | BP | GFR | UA |
| --- | --- | --- | --- | --- | --- | --- | --- | --- | --- | --- |
| P1 | M | 4-exon | c.112C>T | p.Q38* | TAD | nonsense | NA | 146/95 | 162 | - |
| P2 | F | 4-exon | c.273G>A | p.W91* | PRD | nonsense | NA | 120/77 | 90 | - |
| P3 | F | 4-exon |  |  |  | splice* |  | 137/88 | 60 | - |
| P4 | F | 4-exon | c.375G>A | p.T125T | DBD | splice | NA | 111/76 | 59 | + |
| P5 | F | 4-exon | c.375G>A | p.T125T | DBD | splice | NA | 105/80 | 105 | + |
| P6 | F | 4-exon | c.375G>A | p.T125T | DBD | splice | NA | 104/82 | 70 | - |
| P7 | F | 5-exon | c.437G>A | p.W146* | DBD | nonsense | NA | 114/74 | 70 | - |
| P8 | F | 5-exon | c.455C>T | p.P152L | DBD | missense | Del | 122/90 | 125 | + |
| P9 | F | 5-exon | c.455C>T | p.P152L | DBD | missense | Del | 112/69 | 75 | - |
| P10 | M | 5-exon | c.455C>T | p.P152L | DBD | missense | Del | 112/71 | 135 | - |
| P11 | F | 5-exon | c.455C>T | p.P152L | DBD | missense | Del | 115/50 | 150 | - |
| P12 | M | 5-exon | c.455C>T | p.P152L | DBD | missense | Del | 137/80 | 108.5 | - |
| P13 | M | 5-exon | c.473G>A | p.R158H | DBD | missense | Del | 120/80 | 81 | - |
| P14 | F | 5-exon | c.473G>A | p.R158H | DBD | missense | Del | 120/79 | 136 | - |
| P15 | F | 6-exon | c.586C>T | p.R196* | DBD | nonsense | NA | 133/88 | 137.5 | + |
| P16 | F | 6-exon | c.589G>A | p.V197M | DBD | missense | DPF | 99/59 | 66 | - |
| P17 | F | 6-exon | c.637C>T | p.R213* | DBD | nonsense | NA | 145/68 | 90 | - |
| P18 | F | 6-exon | c.659A>G | p.Y220C | DBD | missense | DNF | 140/99 | 90 | - |
| P19 | F | 7-exon | c.733G>A | p.G245S | DBD | missense | DNF | 120/80 | 79 | + |
| P20 | F | 7-exon | c.733G>A | p.G245S | DBD | missense | DNF | 101/70 | 85 | - |
| P21 | F | 7-exon | c.742C>T* | p.R248W | DBD | missense | Del | 105/59 | 122 | + |
| P22 | F | 7-exon | c.743G>A | p.R248Q | DBD | missense | DNF | 139/86 | 109 | - |
| P23 | M | 7-intron | c.783-1G>C |  |  | splice | NA | 130/85 | 126 | - |
| P24 | F | 8-exon | c.844C>T* | p.R282W | DBD | missense | Del | 118/80 | 82 | + |
| P25 | F | 8-exon | c.916C>T | p.R306* | NLS | nonsense | NA | 105/63 | 75 | + |
| P26 | M | 9-intron | c.993+1G>A |  | TD | splice | NA | 156/100 | 220 | - |
| P27 | M |  | DelE1-11 |  |  | deletion |  | 138/90 | 60 |  |
| P28 | M |  | c.376-5_381del11 |  |  | deletion |  | 132/80 | 82 |  |

AA: Amino Acid, PD: Protein Domain, SIFT: Sorting Intolerant From Tolerant, BP: Blood pressure, GFR: glomerular filtration rate, GU: patients with Genitourinary phenotype, + denotes phenotype observed (see table 2 for more details). Del: deleterious, DPF: deleterious/partially functional, DNF: deleterious/non functional, NA: No alteration, \* P3: Splicing variant at codon 125 in exon 4; no additional data available.

**Table 3.** Available information on Genitourinary (GU) phenotype and GFR < 90 of LFS patients reported in this study.

| Patient ID | AA change | BP | GFR | Magnetic Resonance Imaging |
| --- | --- | --- | --- | --- |
| P3 |  | 137/88 | 60 | History of renal and cervical cancers; left kidney, uterus, and ovaries surgically removed. |
| P4 | p.T125T | 111/76 | 59 | Right kidney simple cyst |
| P5 | p.T125T | 105/80 | 105 | Right kidney upper pole: 12 mm lesion consistent with angiomyolipoma |
| P6 | p.T125T | 104/82 | 70 |  |
| P7 | p.W146* | 114/74 | 70 | Breast cancer has been removed |
| P8 | p.P152L | 122/90 | 125 | Right kidney simple cyst measuring 13 x 10 mm. |
| P9 | p.P152L | 112/69 | 75 |  |
| P13 | p.R158H | 120/80 | 81 |  |
| P15 | p.R196* | 133/88 | 137.5 | Breast prosthesis: right kidney mass measuring 10.6 x 8.6 cm, diagnosed as adenoma; gallstones present. |
| P19 | p.G245S | 120/80 | 79 | Right kidney simple cyst. Right kidney 9cm length, left kidney 9.7cm. |
| P20 | p.G245S | 101/70 | 85 | Bilateral mastectomy |
| **P21** | **p.R248W** | **105/59** | **122** | **The right kidney simple cyst measuring 6.8 cm.** |
| **P24** | **p.R282W** | **118/80** | **82** | **Liver and kidney lesions are present. There is a complex renal cyst in the right kidney measuring 3.4 cm.** |
| P25 | p.R306* | 105/63 | 75 | Breast prosthesis and ovarian cysts |
| P27 |  | 138/90 | 60 |  |
| P28 |  | 132/80 | 82 |  |

AA: Amino Acid, BP: Blood pressure, GFR: glomerular filtration rate. Bold indicates the mutations that were further analyzed in this study.

We categorized the variants identified in this cohort based on their genetic mutation type. Six variants contain nonsense mutations leading to premature stop codons and are predicted to result in protein truncation (Lindeboom et al., 2016). Six variants are associated with splicing defects, and one involves a deletion spanning exon 11 within the regulatory domain (Fig. 1). Fifteen are missense variants, all occurring within the DNA-binding domain (DBD; residues 94–292). Many variants associated with kidney phenotypes are located within the DNA-binding domain (DBD). Interestingly, the variants linked to cancer-related phenotypes (IDs 7, 10, 11, 19, and 21) also affect this domain. All these mutations impact the DBD, suggesting that they likely disrupt the structural stability of p53.

**Fig. 1:**
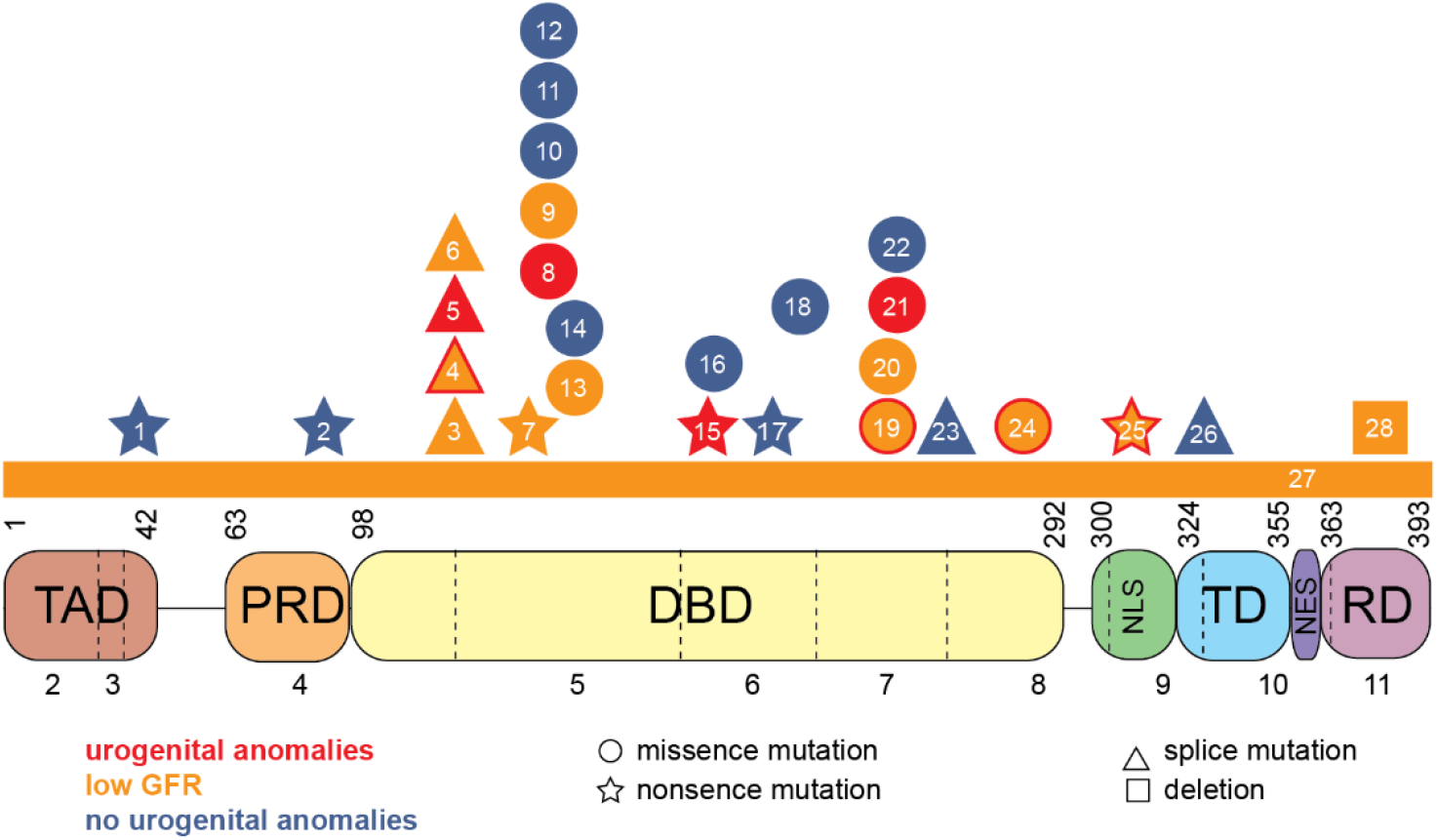
Congenital Anomalies of the Kidney and Urinary Tract (CAKUT) Associated with *TP53* Variants in Li-Fraumeni Syndrome. The schematic illustrates the domain structure of the p53 protein, highlighting patient-specific *TP53* variants. Patient variants are labeled by patient numbers as listed in Table 1. Variants are color-coded as follows: red indicates CAKUT-associated variants; orange marks variants found in patients with low glomerular filtration rate (GFR); orange with a red outline highlights CAKUT-associated variants also associated with low GFR; and blue denotes variants not linked to CAKUT or impaired kidney function. Different shapes indicate the type of variant: a square represents deletion, circles denote missense variants, stars indicate nonsense variants, and triangles mark splice variants. These shapes are positioned according to the affected amino acid sequence. Numbers above the p53 model correspond to amino acid positions, while numbers below indicate exon numbers. Dotted lines represent splice sites. Patients with Li-Fraumeni syndrome exhibit mutations across various p53 domains, including the Trans-Activation Domain (TAD), Proline-Rich Domain (PRD), DNA-Binding Domain (DBD), Nuclear Localization Signal (NLS), Tetramerization Domain (TD), and Regulatory Domain (RD), but not the nuclear export signal (NES). Notably, p53 mutations associated with urogenital anomalies detected by MRI are localized within the DBD.

Of the patients with kidney abnormalities, we selected two distinct human *TP53* variants (identified in patient IDs 21 and 24) for functional modeling in *Xenopus laevis*. These mutations, *TP53*^R248W^ and *TP53*^R282W^, result in missense mutations in the p53 DNA-binding domain and are suspected to exert a dominant-negative effect. Previous studies have indicated that overexpression of a dominant-negative form of *TP53* in *Xenopus* results in the loss of kidney tubules (Wallingford et al., 1997). The *TP53*^R248W^ mutation has been previously characterized, and structural studies of the human p53 core domain revealed that this mutation disrupts DNA binding by eliminating key contacts with the DNA minor groove (Cho et al., 1994). In our studies, the individual carrying this mutation presented with a large kidney cyst measuring 6.8 cm in diameter. The second mutation, *TP53*^R282W^, is novel to this study. Based on its location in the DNA-binding domain and the patient’s clinical features, we hypothesize that it also functions through a similar dominant-negative mechanism. The patient with this mutation exhibited a complex kidney cyst measuring 3.4 cm in diameter, along with additional kidney lesions.

We used AlphaFold to predict structural changes in those variants. p53 functions as a tetramer to bind DNA, and mutant p53 can interact with wild-type p53, inhibiting its function or disrupting DNA binding. The Arginine (Arg =R)-Tryptophan (Trp=W) mutation significantly alters the physicochemical properties of the residue, impacting protein stability and function (Fig. 2A). In the structural prediction, the interaction between guanine/cytosine nucleotides and arginine, hydrophilic and positively charged, is crucial for p53-DNA binding and stabilizes the complex, particularly in the minor groove (Fig. 2B). In contrast, tryptophan is a large, non-polar amino acid that lacks the necessary positive charge, leading to steric hindrance and localized destabilization of the protein. Consequently, the Arg-to-Trp mutation reduces p53’s DNA-binding affinity and disrupts structural integrity (Fig. 2B). Consistent with previous studies, we show that the TP53^R248W^ variant directly affects residues involved in DNA interaction, impairing p53’s ability to bind target gene promoters without significantly altering its overall structure (Roszkowska et al., 2022). On the other hand, the variant residue R282 stabilizes the p53 DNA-binding domain by interacting with neighboring residues such as F134, L130, S127, and Q286 (Fig. 2C). Although it does not directly contact DNA, these interactions are critical for maintaining the protein’s structural integrity. The TP53^R282W^ mutation introduces a bulky tryptophan side chain, causing steric clashes that disrupt this stabilizing network, potentially destabilizing the β-sheet and α-helix interface (Fig. 2C).

**Fig. 2.**
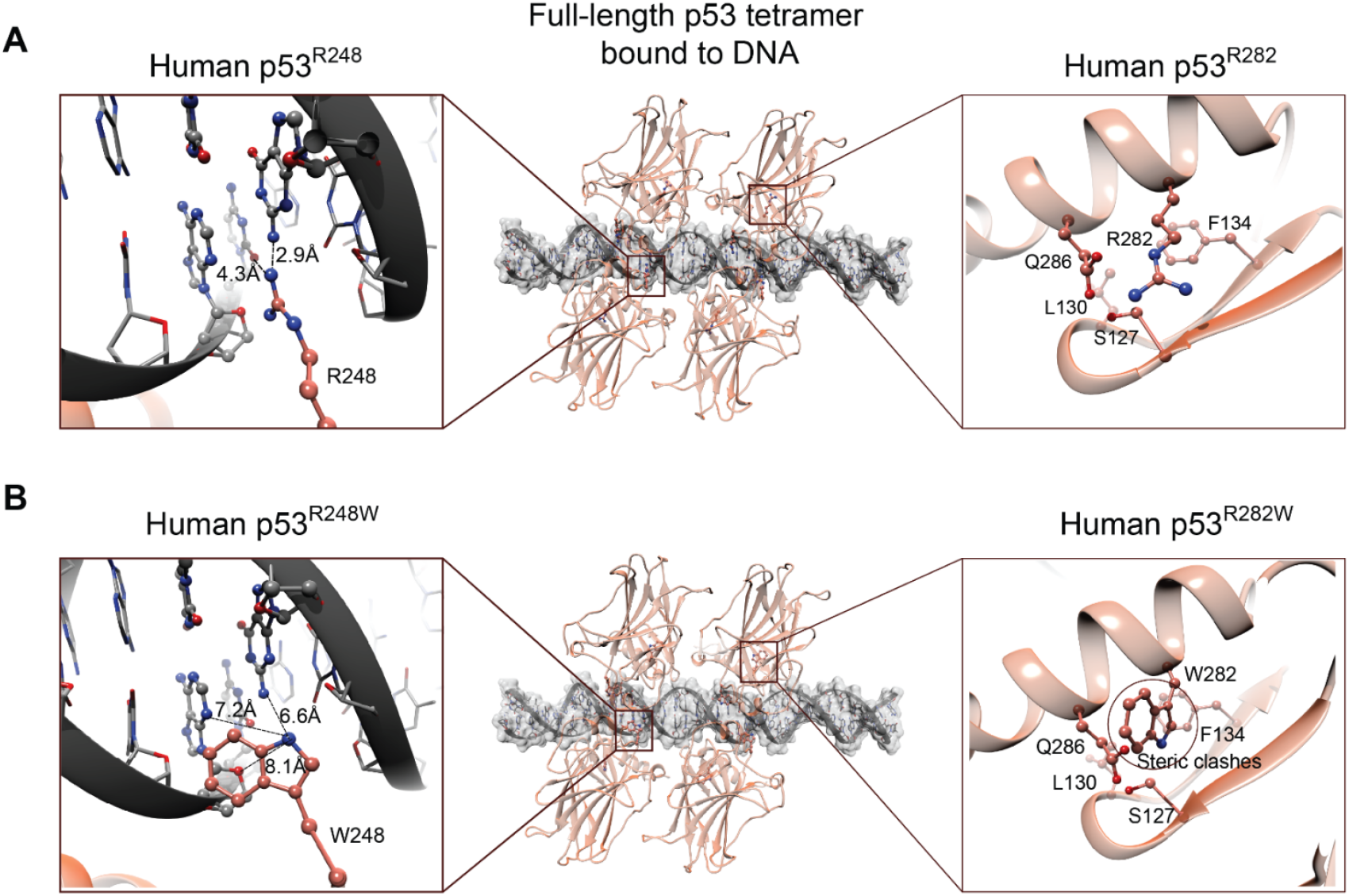
Structural modeling of the p53–DNA complex and effects of p53 R248W and R282W variants. (A) The tetrameric full-length wild-type p53 bound to DNA (center, top). Left: Close-up of residue R248 forming hydrogen bonds with DNA bases (2.9 Å and 4.3 Å). Right: Residue R282 supports structural stability through interactions with neighboring residues (F134, L130, S127, and Q286) but does not directly contact DNA. (B) The tetrameric p53 containing the R248W and R282W mutations bound to DNA (center, bottom). Left: The R248W mutation disrupts hydrogen bonding, increasing distances (6.6–8.1 Å) and likely reducing DNA-binding affinity; the R248 site in the minor groove shows loss of contact. Right: The R282W mutation introduces steric clashes that disrupt the stabilizing network of interactions, potentially destabilizing the β-sheet/α-helix interface of the DNA-binding domain.

### *tp53* is expressed during *Xenopus* development and in the early and late stages of kidney formation

To model human patient data, we used *Xenopus laevis* embryos as a model. To determine if the *X. laevis tp53* gene is homologous to the human, we analyzed the sequence similarity between human and *Xenopus laevis* (Supplementary Information S2). We find that the DNA-binding domain (DBD) is heavily conserved, with 67.34% of the amino acids being identical (Percent Identity Matrix in CLUSTAL O(1.2.4)). As described before, this domain is particularly relevant, as it harbors approximately 80% of *TP53* mutations (Fig. 1) (Bouaoun et al., 2016, Levine, 2019).

To assess endogenous *tp53* expression in the *Xenopus* kidney, *in situ* hybridization utilizing DIG-labeled probes, alkaline phosphatase, and NBT/BCIP was performed at four developmental stages (NF 18, 25, 30, and 40) (Supplementary Information S1A). At stage 18, strong *p53* expression was detected in neuronal structures, including the neural plate, and was widely distributed around the intermediate mesoderm. By stages 25 and 40, *tp53* transcripts were present in the brain, neural tube, head, eyes, and somites (Supplementary Information S1A). During pronephros specification in the early neurula stage (stage 18), *tp53* transcripts were weakly detected but showed a notable reduction by stage 25. However, transcript levels increased again at stage 30, suggesting a potential role in the early epithelialization and differentiation of the pronephros. By stage 40, when the kidney is fully functional and completely epithelialized, *tp53* transcripts were clearly detected in the *Xenopus* kidney (Supplementary Information S1A).

To better visualize *tp53* transcript distribution during kidney development, we employed Hybridization Chain Reaction (HCR) in whole-mount *Xenopus* embryos. This technique uses nucleotide probes that hybridize to target mRNA. The pair of probes contains two initiator sequences that trigger a chain reaction with hairpin molecules (h1 and h2) tagged with fluorescent markers, allowing simultaneous detection of multiple transcripts (Choi et al., 2018). *tp53* transcripts are shown in pink, with *lhx1* and *atp1a1* probes used as markers for nephron progenitors and epithelial kidney tubules, respectively (Fig. 3). Across all stages (18–40, Fig. 3a–f), *tp53* transcripts were highly expressed in neuronal structures, including brain precursors, head, and facial structures. From stages 18–25, *tp53* expression was also detected in the neural crest and neural tube but showed no clear expression in nephron progenitors (Fig. 3a’’–c’’). However, at stages 18 and 22 (Fig. 3a’’ and b’’), *tp53* exhibited a localized pattern around nephron progenitors, suggesting an early role in surrounding cells, possibly in an undifferentiated stage. By stage 25, *tp53* transcripts were no longer detected in the developing kidney or showed only a weak or highly specific signal in kidney progenitors (Fig. 3c’’). At stage 30, *tp53* expression gradually increased, marking a distinct structure around the pronephros (Fig. 3d and d’’). This increase continued, peaking at stage 35, coinciding with tubulogenesis and epithelialization (Fig. 3e). At this stage, *tp53* mRNA levels were markedly elevated in and around the pronephros, with the highest intensity observed in regions surrounding kidney tubules, as marked by *atp1a1* (Fig. 3e and e’’). Atp1a1 is a transporter whose transcript is specifically expressed in epithelial cells of the functional nephron (Fig. 3e’). By stage 40, *tp53* expression was evident inside and around the pronephros (Fig. 3f), with a magnified view suggesting partial colocalization of *tp53* transcripts with kidney markers, reinforcing its potential role in nephron maturation (Fig. 3f and f’’). *tp53* transcript expression is shown throughout the stage 40 tadpole (Fig. 3g). To confirm probe specificity, we performed a negative control by omitting amplifier H2. As expected, using only H1 did not produce any signal, confirming that in situ HCR detection of *tp53* mRNA is specific (Fig. 3h).

**Fig. 3.**
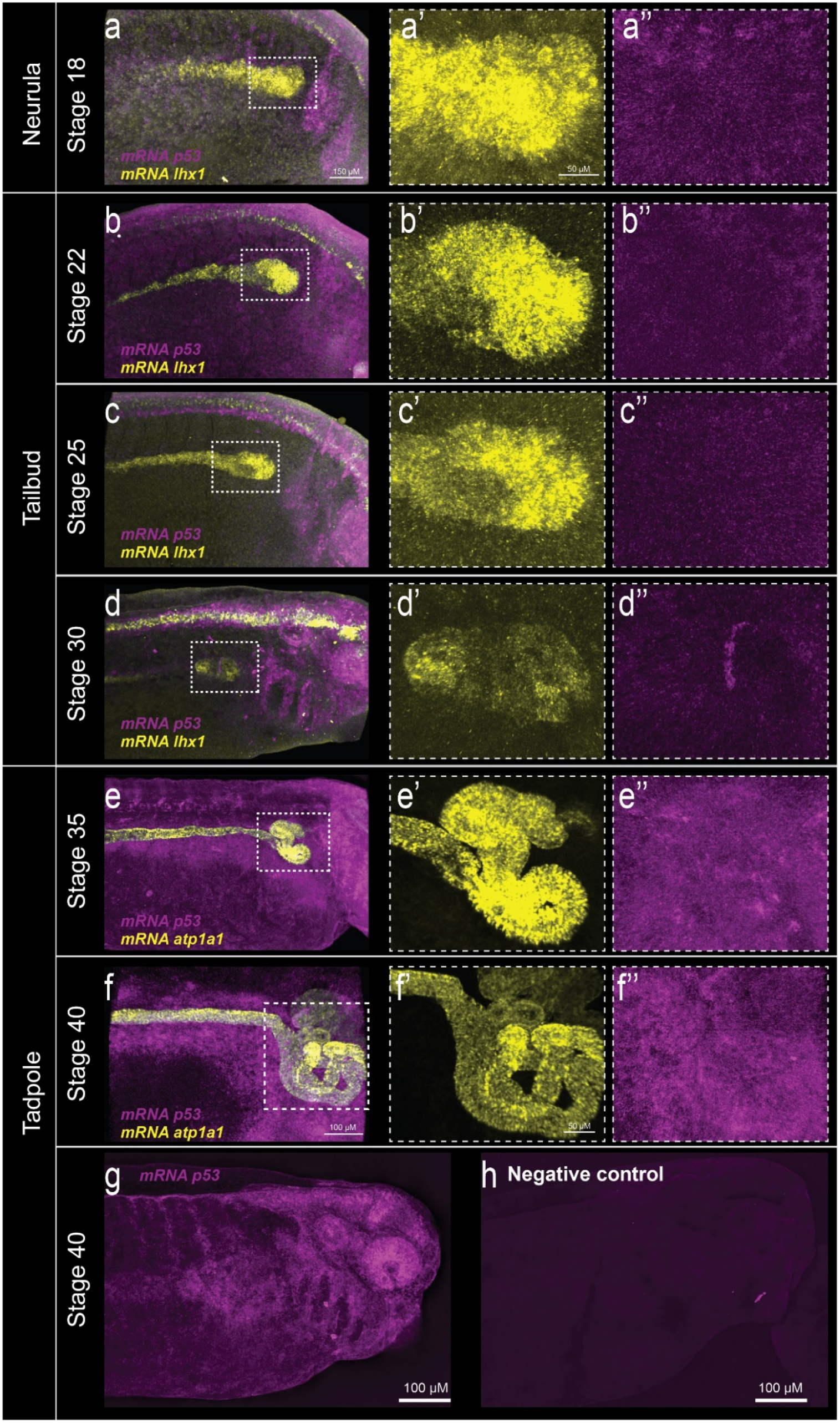
Hybridization Chain Reaction (HCR) whole mount analysis of *tp53* expression during kidney development in *X. laevis*. To examine the spatial–temporal expression of *tp53* mRNA in the kidney, HCR was performed using probes designed against *X. laevis* to target both L and S *tp53* homologous sequences. Pronephric kidney development occurs between stages 12.5 and 40, with nephron progenitor compaction already visible in the neurula stage (stage 18), as indicated by *lhx1* expression (yellow, a-f and a’’-d’’). At stages 18 and 22, p53 transcripts (pink) are detected around, but not within, nephron progenitors (a-b and a’’-b’’). By stage 30, p53 expression increases in specific cells inside and around the differentiating pronephros (d and d’’). However, *tp53* expression is most prominent at stages 35 and 40, coinciding with epithelialization, as marked by *atp1a1* expression (a-f and a’’-f’’). At stage 40, *tp53* is highly expressed both inside and around the differentiated pronephric kidney, with areas of colocalization between *tp53* and *atp1a1* transcripts appearing as white and gray signals (f). A magnified view of the epithelial tubules reveals clear *tp53* expression within kidney tubules (f’ and f’’). Panel a-e show kidneys at stages 18-35 (same 150 µm scale bar). Global *tp53* transcript expression in a stage 40 *X. laevis* embryo. Abundant *tp53* expression is observed in tissues including the facial region, brain, otic vesicle, branchial arches, and kidney. The control, where the harpin H2 was omitted, shows no *tp53* expression, confirming specificity. Each panel (a-e) includes a magnified view of the kidney region (yellow, a’-e’) and *tp53* expression to the right (pink a’’-e’’). Scale bars: 50 µm (a’-f’ and a’’-f’’) and 100 µm (stage 40, f).

Western blot analysis on whole embryo lysates confirmed p53 protein expression throughout the stages of kidney development, showing a marked increase after stage 30 (tailbud) and peaking at stages 35 and 40 (Supplementary Information S1B). The p53 Western blot consistently displays two bands. The lower band runs at the size seen for other organisms, but it is unclear whether the upper band is a modified protein or nonspecific binding (Supplementary Information S1B).

### Missense *TP53* variants R248W and R248W from Li-Fraumeni patients lead to abnormal kidney development in *Xenopus*

Site-directed mutagenesis was used to introduce mutations, and mRNA for *TP53*^R248W^, *TP53*^R282W^, and wild-type human *TP53* was synthesized. All constructs contained two FLAG epitopes. Western blot analysis with TP53- and FLAG-specific antibodies confirmed the successful expression of the proteins derived from wild-type *TP53, TP53*^R248W^, and *TP53*^R282W^ transcripts in *Xenopus* embryos (Fig. 4F). Notably, *TP53*^R282W^ exhibited lower expression levels than wild-type *TP53* and *TP53*^R248W^ (Fig. 4), likely due to reduced protein stability resulting from impaired conformational integrity. To evaluate the functional impact of these mutations, we injected wild-type *TP53, TP53*^R248W^, or *TP53*^R282W^ mRNA into the V2 blastomere of 8-cell *Xenopus* embryos to target a single kidney. A subphenotypic dose of wild-type *TP53* was established to prevent aberrant kidney development, as high p53 levels have been previously shown to induce defects (Hoever et al., 1994). Injection of 300 pg of *TP53*^R248W^ or *TP53*^R282W^ mRNA significantly reduced kidney tubule formation (Fig. 4C and D), whereas wild-type *TP53* had no such effect (Fig. 4B), indicating a dominant negative effect of the patient variants on nephric tubule formation.

**Fig. 4:**
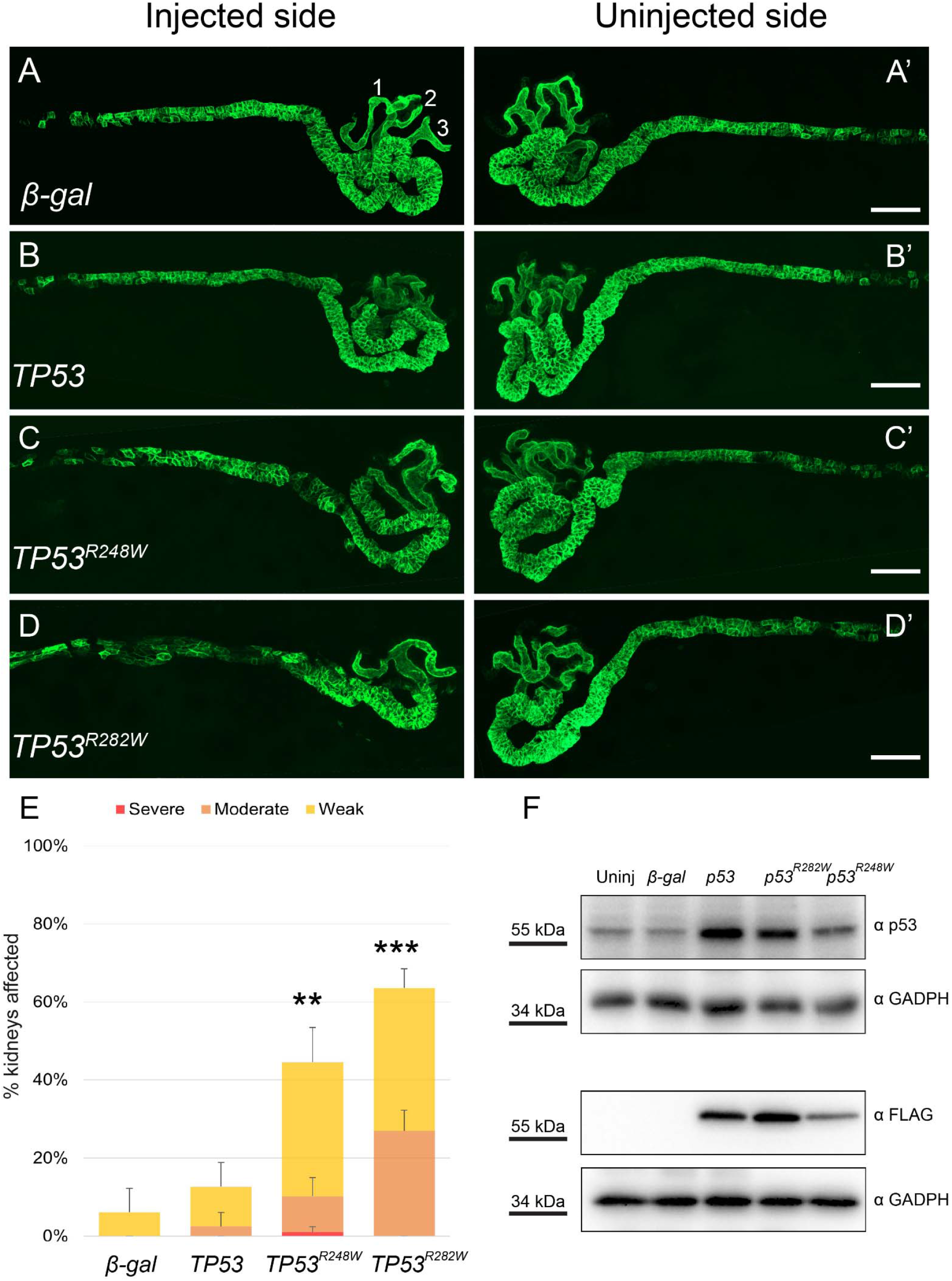
Mutant Human *TP53* Impairs Kidney Development in *Xenopus* Embryos. (A–D) *Xenopus* embryos were unilaterally injected at the 8-cell stage with 300 pg of β*-galactosidase (*β*-gal)*, wild-type human *TP53, TP53*^*R248W*^, or *TP53*^*R282W*^ mRNA. At stage 40, tadpoles were stained with kidney-specific antibodies: 3G8 (proximal tubules) and 4A6 (distal and connecting tubules). Panels without apostrophes (A–D) depict the injected side, whereas corresponding panels with apostrophes (A’–D’) show the uninjected side. Overexpression of mutant p53 (C–D) disrupted kidney development, whereas wild-type p53 (B) had no effect. β-gal (A) served as a negative control. Scale bars represent 100 μm. (E) Quantification reveals a significant reduction in kidney tubule formation in embryos injected with *TP53*^*R248W*^ or *TP53*^*R282W*^ mRNA compared to those injected with wild-type p53. Kidney phenotypes were assessed at Nieuwkoop and Faber stage 40 (Nieuwkoop and Faber, 1994). A normal kidney exhibits three proximal branches connected to a central trunk (showed with numbers in A), leading to the convoluted early distal region and strong immunostaining in the late distal region. A **weak** phenotype is characterized by the loss of one proximal branch, reduced early distal looping, and/or partial loss of late distal immunostaining. A **moderate** phenotype involves the loss of two proximal branches, moderate reduction in early distal looping, and potential late distal immunostaining loss. A **severe** phenotype is defined by the complete absence of proximal tubules, along with severe or total loss of early distal looping and late distal structures. Phenotypes examples for this nomenclature were published by (Blackburn et al., 2019). Statistical significance was assessed using a two-tailed *t*-test, with weak (yellow bar), moderate (orange bar), and severe (red bar) kidney phenotypes. Double asterisks (**) denote *p* < 0.007, and triple asterisks (***) denote *p* < 0.0006. Error bars represent the standard error. (F) Single-cell *Xenopus* embryos were injected with 1 ng of wild-type, R248W, or R282W human *TP53* mRNA alongside a membrane RFP tracer. Western blot analysis confirmed successful translation of wild-type and mutant *TP53* mRNA in *Xenopus* embryos. Notably, the R282W mutant exhibited lower expression levels compared to wild-type *TP53* and the R248W mutant, suggesting reduced protein stability. β-gal was used as a negative control. The R248W (C742T) and R282W (C844T) point mutations were introduced via site-directed mutagenesis using a wild-type human *pcDNA3-FLAG-p53* vector. GAPDH served as a loading control, and molecular weight markers are indicated on the left side of the blot.

## Discussion

Our findings contribute to a growing body of evidence implicating *TP53* mutations, particularly those with dominant-negative properties, in congenital anomalies of the kidney and urinary tract (CAKUT) within the context of Li-Fraumeni syndrome (LFS). Although p53 is classically characterized as a tumor suppressor, accumulating data now indicate its broader involvement in embryonic development, including roles in nephrogenesis and organ patterning (Li et al., 2015, Saifudeen et al., 2009). This study uncovers overlooked developmental roles of p53 mutations and expands the phenotypic spectrum of Li-Fraumeni syndrome to include urogenital anomalies.

The identification of CAKUT, including kidney hypoplasia, renal agenesis, and/or reduced glomerular filtration rates in approximately one-third of LFS patients, suggests a non-random association between p53 dysfunction and renal developmental defects. The prevalence of impaired renal function (GFR <90) in nearly half of the cohort further supports a physiologically relevant role for p53 in renal morphogenesis. Importantly, our functional experiments in *Xenopus laevis* confirmed that the overexpression of two human *TP53* variants, identified in LFS patients, causes the developmental disruptions in nephrogenesis similar to those observed with p53 loss (Saifudeen et al., 2002, Saifudeen et al., 2009), implicating both loss-of-function and/or dominant-negative mechanisms. Notably, specific hotspot mutations R248W and R282W are strongly associated with kidney phenotypes in the patient cohort of this study. These mutations occur in the highly conserved DNA-binding domain of p53 and have distinct structural and functional consequences. R248W, a DNA-interaction mutation, has previously been shown to aberrantly activate oncogenic pathways, including STAT3 signaling (Klemke et al., 2021, Schulz-Heddergott et al., 2018). Its correlation with kidney phenotypes in both human patients and *Xenopus laevis* model suggests that this variant may interfere with p53’s transcriptional regulation of developmental genes. Similarly, R282W destabilizes the DBD by perturbing the loop-sheet-helix motif, potentially leading to protein misfolding and compromised DNA-binding (Joerger et al., 2006). These mechanistic insights suggest that certain p53 mutations may selectively disrupt developmental pathways independently of their tumor-suppressive roles, though further studies are needed to clarify this.

The evolutionary conservation of p53 across vertebrates and non-vertebrates underscores its critical role beyond cancer biology (Brandt et al., 2009, Armstrong et al., 1995). The observation that *Xenopus* (a non-amniote) develops kidney abnormalities upon expression of human *TP53* mutants underscores the conserved, evolutionary critical role of p53 in organogenesis. Our results highlight further consideration of the wider developmental consequences of p53 dysfunction. While p53 activity is tightly regulated to prevent inappropriate apoptosis or senescence during embryogenesis (Molchadsky et al., 2010), our data suggest that aberrant p53 signaling can disrupt organ development. This is particularly relevant in LFS, where germline *TP53* mutations may predispose individuals not only to malignancy but also to developmental abnormalities, including CAKUT.

In conclusion, this study provides novel evidence that p53 plays a vital role in renal development and that specific *TP53* variants associated with LFS can contribute to CAKUT. These findings open new avenues for exploring p53’s developmental functions and underscore the importance of considering non-oncogenic phenotypes in patients with germline *TP53* mutations. Future studies should focus on identifying downstream p53 targets involved in nephrogenesis and delineating the molecular pathways disrupted by specific p53 variants during embryonic development.

## Data Availability

All data produced in the present work are contained in the manuscript.

## Acknowledgements

We sincerely thank the patients and their families for their participation in this study. The SIGNIFY study was funded by The Annabel Evens Memorial Fund and supported by the National Institute for Health and Care Research (NIHR) to the Biomedical Research Centre at The Royal Marsden NHS Foundation Trust and Institute of Cancer Research. We are grateful to the faculty and staff of the Royal Marsden NHS Foundation Trust, London, UK and Central Manchester University Hospitals NHS Foundation Trust, Manchester, UK for their contributions. We acknowledge the SIGNIFY Study Steering Committee: Prof Anwar Padhani, Paul Strickland Scanner Centre, Mount Vernon Cancer Centre; Prof Leslie Walker, University of Hull; Dr Gillian Mitchell, Familial Cancer Centre, Peter MacCallum Cancer Centre and Sir Peter MacCallum Department of Oncology, University of Melbourne; Dr Gek Kwan-Lim, The Institute of Cancer Research and The Royal Marsden NHS Foundation Trust; Susan Eastbrook, patient representative; Dr Peter Simmonds, Cancer Sciences Academic Unit and University of Southampton Clinical Trials Unit, Faculty of Medicine, University of Southampton and University Hospital Southampton Foundation Trust; Dr Frank Saran, The Royal Marsden NHS Foundation Trust. We also appreciate the helpful input from members of the laboratories of Drs. Rachel K. Miller and Rosalind A. Eeles. We thank the instructors and teaching assistants of the Cold Spring Harbor Laboratory’s Cell & Developmental Biology of *Xenopus*: Gene Discovery & Disease Course for advanced research training, particularly Drs. Lance A. Davidson and Chenbei Chang, who directed the course in 2022-2024. Particular thanks to Dr. Carole LaBonne and Andrew Montequin for their assistance with the HCR protocol. This work was supported by the NIDDK (R01DK115655 to R.K.M.), and *Xenopus* studies were funded by K01DK092320 and R03DK118771 (to R.K.M.), as well as startup funds from the Department of Pediatrics, Pediatric Research Center, McGovern Medical School. We thank members of the Miller and McCrea labs, and Dr. M. Kloc, for their valuable suggestions. Special thanks to Dr. T.H. Gomez for animal care. Confocal imaging was supported by the UTHealth Office of the Executive Vice President and Chief Academic Officer and the Department of Pediatrics Microscopy Core through the Zeiss LSM800 confocal microscope.

## Disclosure of Interests

Professor Rosalind Eeles has the following conflicts of interest to declare: Honoraria from GU-ASCO, Janssen, University of Chicago, Dana Farber Cancer Institute USA as a speaker. Educational honorarium from Bayer and Ipsen, member of external expert committee to Astra Zeneca UK and Member of Active Surveillance Movember Committee. She is a member of the SAB of Our Future Health. She undertakes private practice as a sole trader at The Royal Marsden NHS Foundation Trust and 90 Sloane Street SW1X 9PQ and 280 Kings Road SW3 4NX, London, UK.

## Notes

### Funding Statement

This study was funded by the National Institute for Health and Care Research (NIHR) to the Biomedical Research Centre at The Royal Marsden NHS Foundation Trust and Institute of Cancer Research.
The Xenopus work was supported by the NIDDK (R01DK115655 to R.K.M.) and by K01DK092320 and R03DK118771 (to R.K.M.).

### Author Declarations

The subjects that were recruited and consented to enrollment in the study in accordance with an institutional IRB at the The Institute for Cancer Research, as described in Saya et al. Familial Cancer 2017. The research was approved by the Health Research Authority NRES Committee London-Brent (12/LO/0781).

