## Supplementary Information for "Li-Fraumeni Syndrome-Associated p53 Variants Disrupt Kidney and Urinary Tract Development"

Fig. S1

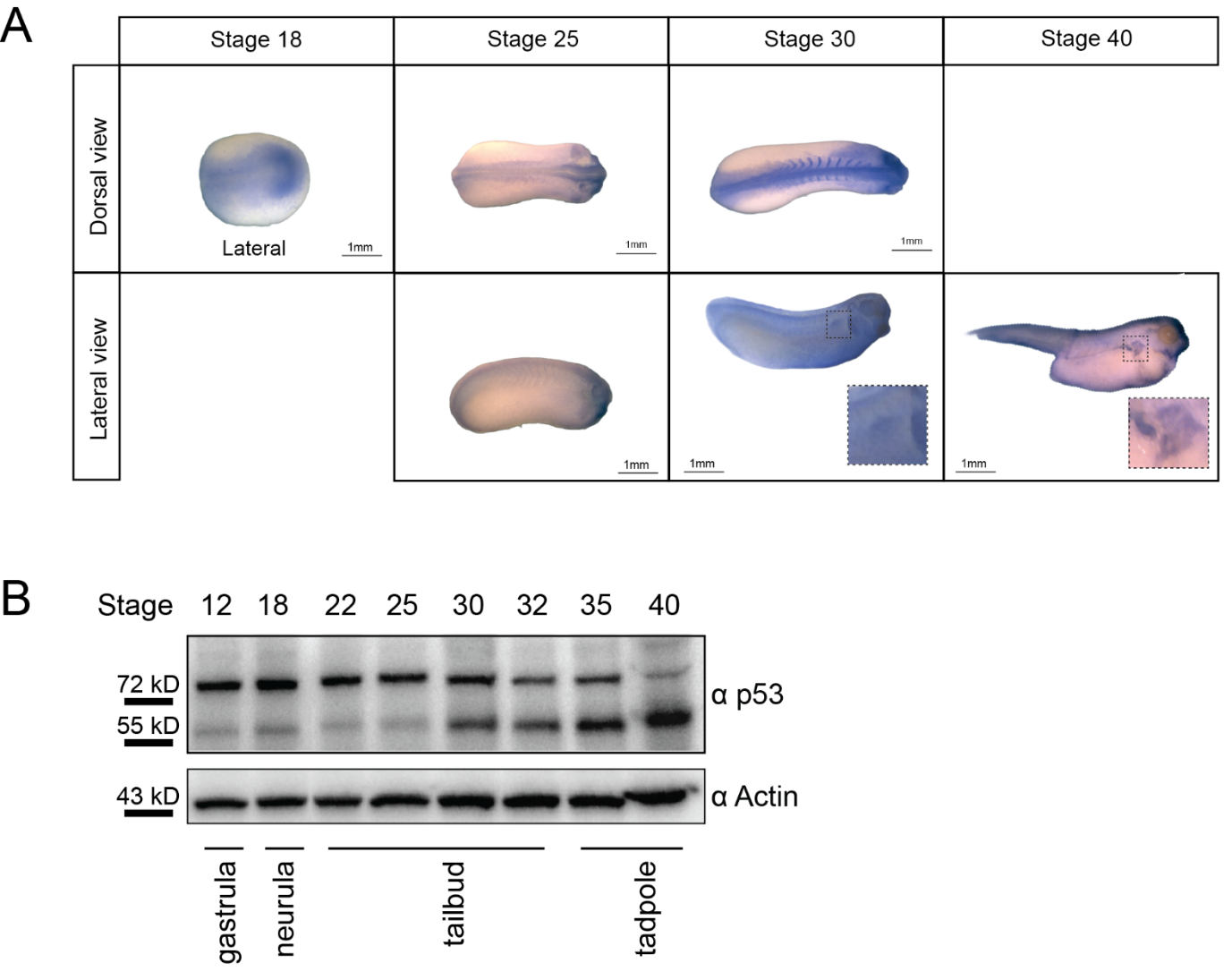

**Fig. S1: p53 expression across developmental stages.** (A) *In situ* hybridization of *tp53* at various developmental stages shows widespread expression in the central nervous system of *X. laevis*, with specific expression observed in the pronephric kidney at stage 30. (B) p53 Western blot analysis across developmental stages. Protein extracts from *X. laevis* embryos at the indicated Nieuwkoop and Faber stages were prepared and subjected to Western blot analysis. Actin (Sigma A2066) was used as a loading control, while p53 was detected using the p53 [X77] antibody from Abcam.

Fig. S2

```
sp|P07193|P53_XENLA      -MEPSSETGMDPPLSQETFEDLWSLLPDPLQTVTCRL---DNL-------SEF-------	42
sp|P04637|P53_HUMAN      MEEPQSDPSVEPPLSQETFSDLWKLLPENNVLSPLPSQAMDDLMLSPDDIEQWFTEDPGP	60
                           **.*: .::********.***.***:            *:*       .::

sp|P07193|P53_XENLA      ---PDYPLAADMTVL-----QEGLMGNAVPTVTSCAVPSTDDYAGKYGLQLDFQQNGTAK	94
sp|P04637|P53_HUMAN      DEAPRMPEAAPPVAPAPAAPTPAAPAPAPSWPLSSSVPSQKTYQGSYGFRLGFLHSGTAK	120
                            *  * **  ..        .  . *     *.:*** . * *.**::*.* :.****

sp|P07193|P53_XENLA      SVTCTYSPELNKLFCQLAKTCPLLVRVESPPPRGSILRATAVYKKSEHVAEVVKRCPHHE	154
sp|P04637|P53_HUMAN      SVTCTYSPALNKMFCQLAKTCPVQLWVDSTPPPGTRVRAMAIYKQSQHMTEVVRRCPHHE	180
                         ******** ***:*********: : *:* ** *: :** *:**:*:*::***:******

sp|P07193|P53_XENLA      RSVEPGEDAAPPSHLMRVEGNLQAYYMEDVNSGRHSVCVPYEGPQVGTECTTVLYNYMCN	214
sp|P04637|P53_HUMAN      RCSD-SDGLAPPQHLIRVEGNLRVEYLDDRNTFRHSVVVPYEPPEVGSDCTTIHYNYMCN	239
                         *. : .:. ***.**:******:. *::* *: **** **** *:**::***: ******

sp|P07193|P53_XENLA      SSCMGGMNRRPILTIITLETPQGLLLGRRCFEVRVCACPGRDRRTEEDNYTKKRGLK---	271
sp|P04637|P53_HUMAN      SSCMGGMNRRPILTIITLEDSSGNLLGRNSFEVRVCACPGRDRRTEEENLRKKGEPHHEL	299
                         *******************  .* ****..*****************:*  **   :

sp|P07193|P53_XENLA      PSG--KRELAHPPSSEPPLPKKRLVVVDDDEEIFTLRIKGRSRYEMIKKLNDALELQESL	329
sp|P04637|P53_HUMAN      PPGSTKRALPNNT-SSSPQPKKK----PLDGEYFTLQIRGRERFEMFRELNEALELKDAQ	354
                         * *  ** * :   *. * ***:      * * ***:*:**.*:**:::**:****:::

sp|P07193|P53_XENLA      DQQKVTIKCRKCRDEI------KPKKGKKLLVKDEQPDSE	363
sp|P04637|P53_HUMAN      AGKEPG-GSRAHSSHLKSKKGQSTSRHKKLMFKTEGPDSD	393
                           ::    .*   ..:      . .: ***:.* * ***:
```

**Fig. S2: Sequence alignment of p53 in *Xenopus laevis* and human.** The alignment illustrates conservation across amino acid residues. The protein p53 Human DNA-binding domain (DBD) is highlighted in gray (residues 94–292). Residues corresponding to newly identified variants in this study are marked in green; the two variants modeled in *Xenopus laevis* in this study are indicated in yellow and red. Symbols denote conservation as follows: * = Fully conserved residues, **:** = Conservation between groups of strongly similar properties (Gonnet PAM 250 score >0.5), **.** = Conservation between groups of weakly similar properties (Gonnet PAM 250 score ≤0.5), (no symbol) = Non-conserved residues. Percent Identity Matrix = 54.49%. Data from CLUSTAL O(1.2.4) multiple sequence alignment.

Table S1

| Average | | | | | | |
| --- | --- | --- | --- | --- | --- | --- |
|  | *β-gal* | *TP53* | | *R248W* | *R282W* | |
| Weak | 6.11 | 10.13 | | 34.36 | 36.74 | |
| Moderate | 0.00 | 2.50 | | 9.14 | 26.87 | |
| Severe | 0.00 | 0.00 | | 1.00 | 0.00 | |
| Standard Error | | | | | | |
| Weak | 6.06 | 6.21 | | 8.98 | | 4.81 |
| Moderate | 0.00 | 3.54 | | 4.79 | | 5.28 |
| Severe | 0.00 | 0.00 | | 1.41 | | 0.00 |
| Biological replicates | | | | | | |
|  | 1 | 2 | | 3 | | 4 |
| *β-gal* | 0.00 | 18.18 | | 0.00 | | 6.25 |
| *TP53* | 21.43 | 9.09 | | 0.00 | | 20.00 |
| *R248W* | 50.00 | 28.00 | | 42.86 | | 57.14 |
| *R282W* | 77.78 | 60.00 | | 50.00 | | 66.67 |
| **t-test** | | | | | | |
| *β-gal* vs *TP53* | | | 0.361853 | | | |
| *β-gal* vs *TP53 R248W* | | | 0.002266 | | | |
| *β-gal* vs *TP53 R282W* | | | 0.000212 | | | |
| *TP53 vs R248W* | | | 0.00727 | | | |
| *TP53 vs R282W* | | | 0.000573 | | | |

**Table S1: Statistical analysis.** Summary of the analysis results and significance used to generate Figure 4E.The average, standard error, and totals from four biological replicates are shown as percentages. Statistical significance was assessed using a two-tailed t-test. The total number of *Xenopus* embryos analyzed for each condition was as follows: *β-gal* = 43, *TP53* = 39, *R248W* = 53, and *R282W* = 63.
